# Outbreak of *Neisseria gonorrhoeae* ST16676 among disseminated infections in Minnesota, USA, 2025

**DOI:** 10.64898/2026.01.09.26343522

**Authors:** Daniel Evans, Allison LaPointe, Christine Peel, Khalid Bo-Subait, Elizabeth Dufort, Jenell Stewart, John Kaiyalethe, Bradley Craft, Matthew Plumb, Bonnie Weber, Laura Bohnker-Voels, Kelly Pung, Alyssa Mondelli, Jacob Garfin, Sarah Namugenyi, Paula Snippes-Vagnone, M. Elizabeth Gyllstrom, Kathryn Como-Sabetti, Ruth Lynfield

## Abstract

We summarize an outbreak of *N. gonorrhoeae* ST16676 associated with disseminated gonococcal infections (DGIs) in Minnesota, USA in 2025. This novel strain replaced ST11184 as the predominant sequence type circulating among DGI cases in the state, encoded a *porB1a* allele, and carried a tetracycline resistance gene on a mobilizable plasmid.

## INTRODUCTION

The sexually transmitted pathogen *Neisseria gonorrhoeae* can circulate from mucosal tissue at sites of exposure to other locations in the body, causing disseminated gonococcal infections (DGIs) [1]. After DGI cases increased nearly four-fold in 2024, the Minnesota Department of Health (MDH) expanded DGI surveillance to include whole-genome sequencing (WGS) analysis of isolates [2]. We continued DGI genomic surveillance during 2025 to continue classifying strains and track suspected outbreaks.

## THE INVESTIGATION

In 2025, cases of DGI continued to occur at an elevated incidence rate compared to the 2020-2023 baseline. From January through September 2025, 28 verified DGI cases with detection of *N. gonorrhoeae* from disseminated sites of infection were reported by Minnesota healthcare facilities to MDH. Minnesota state reporting rules that require *N. gonorrhoeae* specimens from normally sterile sites be submitted to the state public health laboratory. Analysis of these cases and linked specimens is considered enhanced surveillance and therefore deemed exempt from Institutional Review Board approval.

We confirmed the species and constructed WGS libraries from 28 *N. gonorrhoeae* isolates using the same protocols as previously described [2]. We performed whole-genome sequencing using the Illumina MiSeq, NextSeq, or MiSeq i100 platforms (Illumina, Inc.) and assembled high-quality genomes of 27 (96.4%) isolates. We identified multi-locus sequence types (MLSTs), *N. gonorrhoeae* sequence types by antimicrobial resistance (NG-STAR), porin B allele types, and gonococcal genetic island (GGI) sequences using previously described approaches [3–6]. We then constructed a midpoint-rooted, core genome phylogenetic tree of all DGI genomes through September 2025 using Bakta v1.9.4, Panaroo v1.5.0, and IQTree2 v2.3.6 (Figure 1) [7–9].

**Figure 1:**
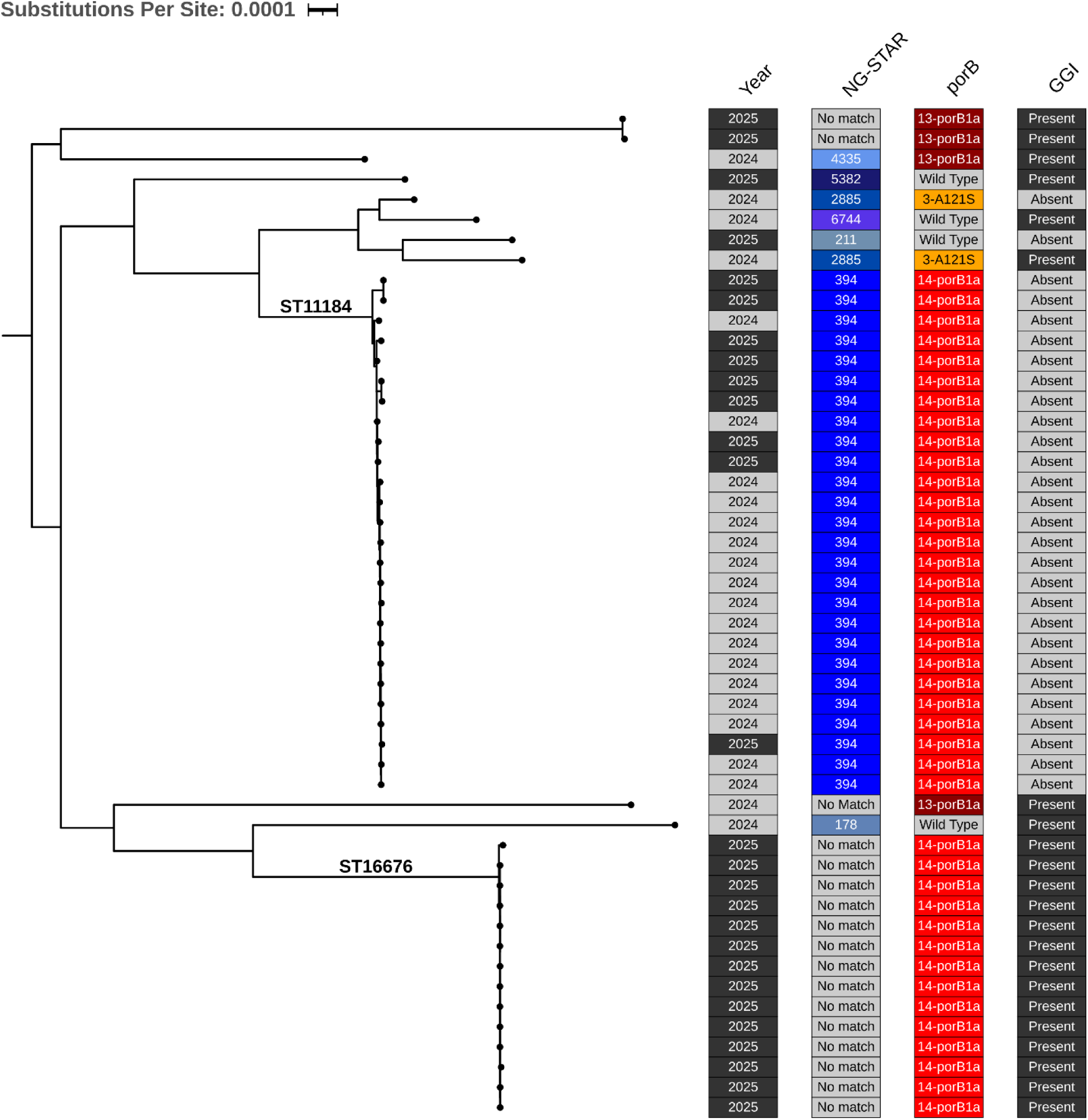
Midpoint-rooted phylogenetic tree constructed from an alignment of 1626 core genes shared by 50 genomes of *N. gonorrhoeae* isolates from disseminated infections in Minnesota in 2024 and 2025. Annotations denote calendar year of specimen collection, NG-STAR, porB allele type, and presence of a gonococcal genetic island (GGI). Clades of genomes assigned to ST16676 and ST11184 are labeled on the tree. This figure was constructed using ITOL software (https://itol.embl.de/).

Our genomic surveillance showed that a new MLST, ST16676, replaced ST11184 as the predominant MLST among 2025 DGI cases (Figure 1, Figure 2). Of the 22 sequenced isolates from specimens collected from June through September 2025, 14 (63.6%) belonged to this MLST (Figure 2). ST16676 genomes did not match any documented NG-STAR profiles, carried a GGI, and encoded the same DGI risk-associated *porB1a* allele present in the ST11184 strain [2, 10].

**Figure 2:**
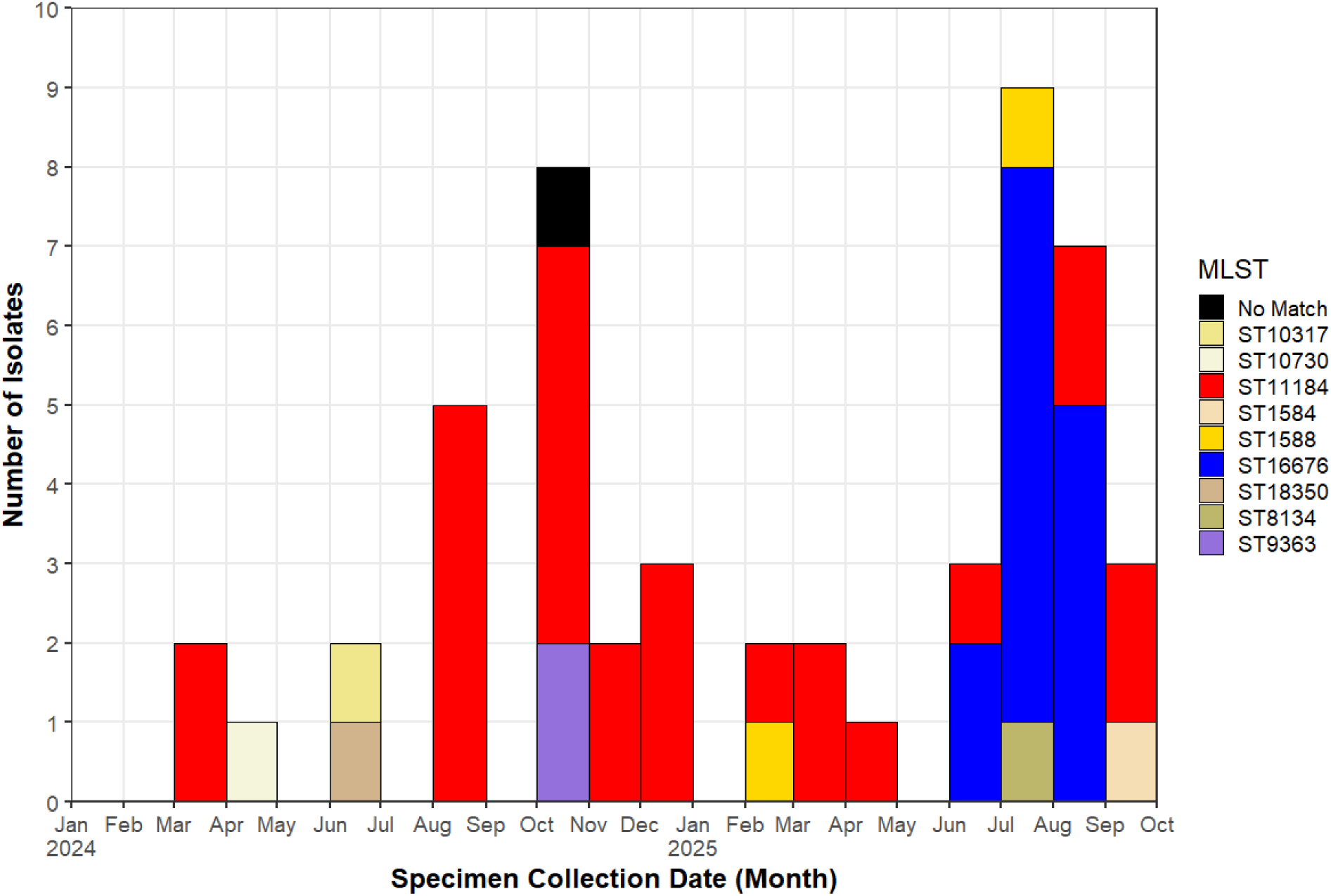
Epidemic curve of Minnesota DGI cases in 2024 and 2025. Cases are binned by calendar month of specimen collection and annotated by multi-locus sequence types (MLSTs) of sequenced isolates.

We searched genomes for markers of antimicrobial resistance (AMR) with AMRFinderPlus v4.0.23 and by examining their NG-STAR alleles [6, 11]. Every ST16676 genome carried the tetracycline resistance gene *tet(M)*, the extended spectrum beta-lactamase gene *bla_TEM_*, and a Type XIV non-mosaic *penA* allele (Appendix Figure 1, Table 1). To characterize AMR-encoding loci, we performed long-read sequencing of four ST16676 isolates using the Rabit Barcoding Kit 24 V14 (SQK-RBK114.24) and R10.4.1 sequencing chemistry on the GridION platform (Oxford Nanopore Technologies). Hybrid assemblies constructed with Unicycler v0.5.0 consistently resolved the acquired *tet(M)* and *bla_TEM_* genes on separate plasmid sequences of 42kb and 5.6kb and the *porB1a* allele on the bacterial chromosome [12].

**Table 1:**
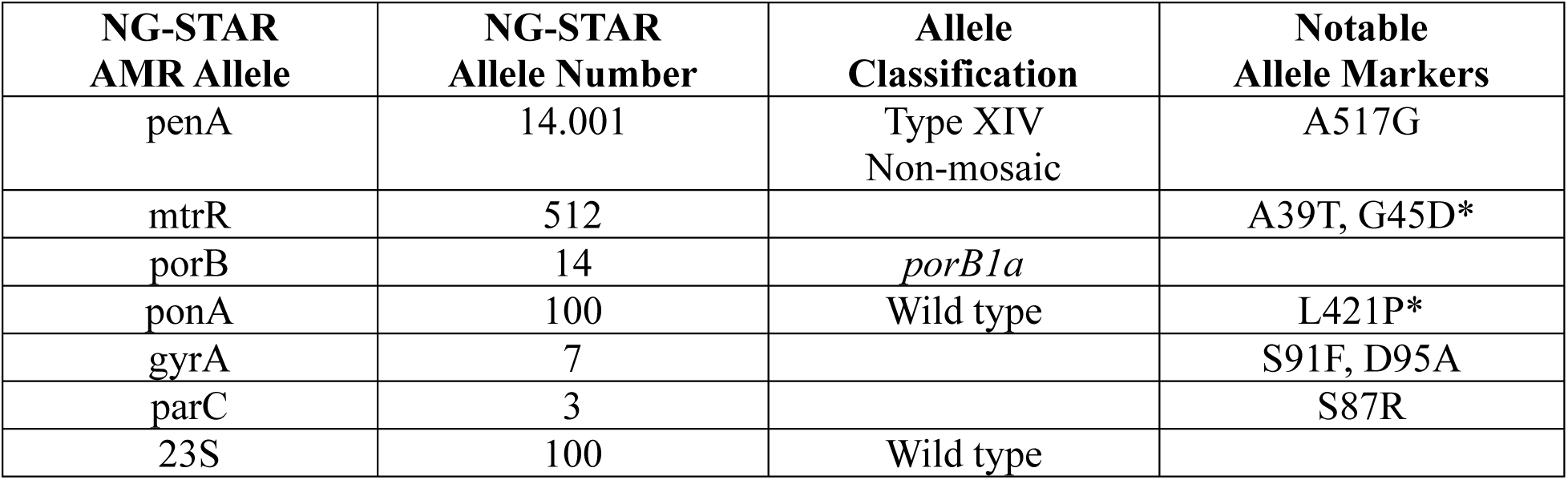
Summary of sequence typing by antimicrobial resistance (NG-STAR) alleles carried by the ST16676 strain. Asterisks denote AMR-associated point mutations that AMRFinderPlus detected in the gene sequences housing those alleles but did not influence NG-STAR classifications.

To determine whether the ST16676 strain was unique to Minnesota, in October 2025, we uploaded genomic data to the National Center for Biotechnology Information (NCBI) database for global comparisons to other genomes in its Pathogen Detection genomic surveillance pipeline. That month, Pathogen Detection grouped the 14 DGI isolates’ genomes into a SNP cluster (PDS000214546.4) with 12 other genomes, which had specimen collection dates between November 2023 and December 2024. We downloaded publicly available sequencing reads for these 12 genomes and assessed their relatedness to the 14 DGI genomes with the Dryad v3.0 pipeline, using the earliest 2025 DGI genome as an internal reference [13]. This approach also divided the Minnesota and non-Minnesota genomes into two separate clades (Figure 3, Appendix Figure 1). Minnesota DGI genomes ranged in genetic similarity to each other by 0 to 62 single nucleotide polymorphisms (SNPs, median 6 SNPs) – with 13 of the 14 ranging from 0 to 16 SNPs (median 6 SNPs) – and to the other 12 genomes by 215 to 320 SNPs (median 248 SNPs). All non-Minnesota genomes were assigned to ST16676, but they lacked *porB1a* alleles and were assigned to a different NG-STAR classification (5262).

**Figure 3:**
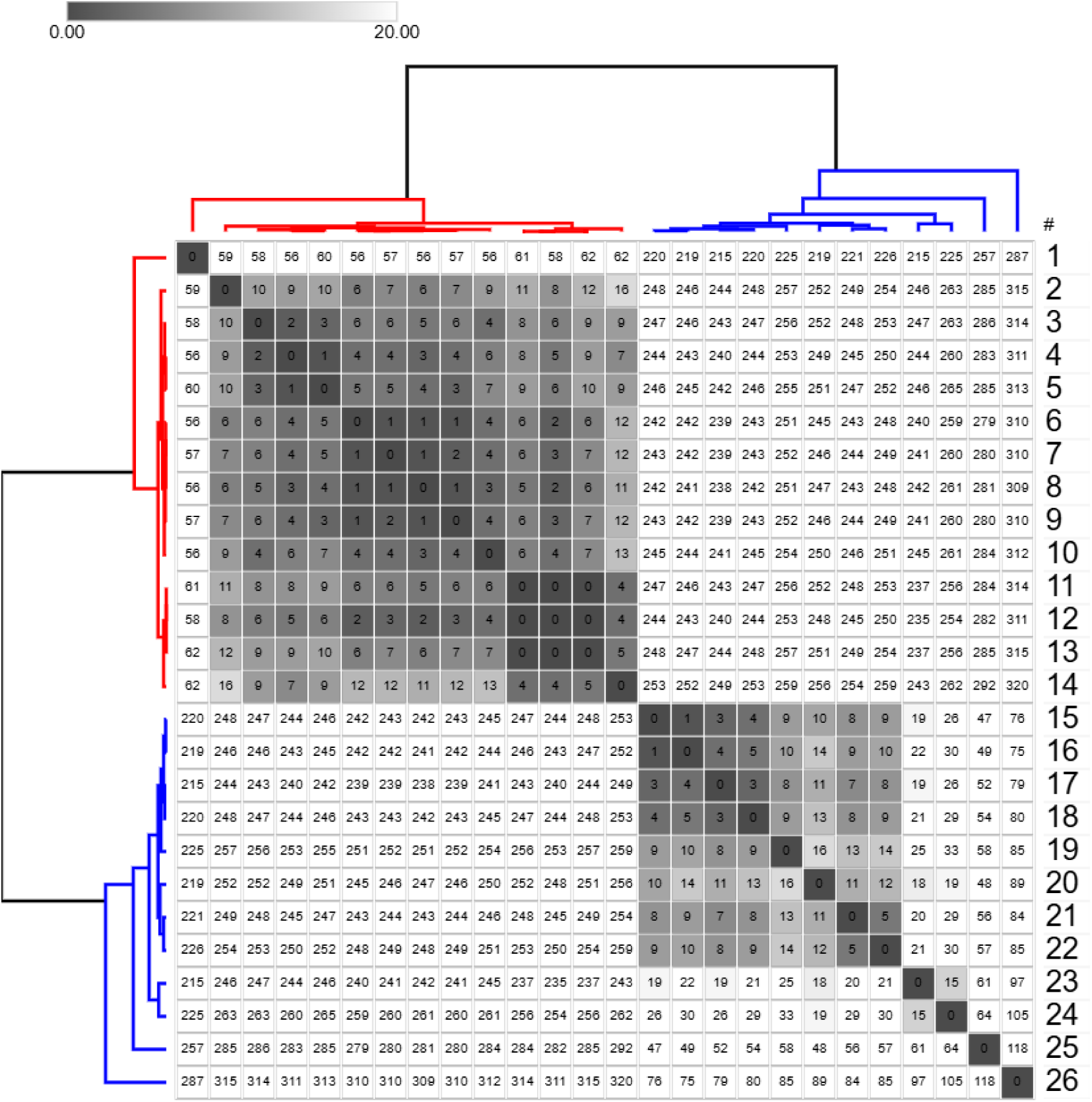
Reference-based pairwise SNP matrix of 26 *N. gonorrhoeae* ST16676 infections. Genomes numbered 1-14, highlighted in red, are from 2025 DGI isolates, and genomes numbered 15-26, highlighted in blue, are the other genomes in the same Pathogen Detection cluster. SNP calls were clustered and displayed using Morpheus software (Broad Institute, Cambridge, Massachusetts).SNP calls less than 20 are highlighted in grayscale. Genome number 7 was used as an internal reference for calling SNPs with the Dryad v3.0 pipeline.

To estimate when the Minnesota ST16676 strain might have emerged, we constructed a core genome alignment and phylogenetic tree of all 26 ST16676 genomes and then used TreeTime v0.11.4 to perform a phylodynamic analysis (Appendix Methods, Appendix Figure 1) [14]. Of the 32 iterations of this analysis that we performed (median R^2^ of molecular clock models = 0.92) (Appendix Methods), the outputs of 28 – including all models whose clock models yielded R^2^ values greater than 0.8 – converged in early May 2025 as an estimated time of a most recent common ancestor (T_MRCA_) for all 14 Minnesota genomes.

ST11184 strains carrying *porB1a* alleles predominated during 2024 and before June 2025, and they continued to persist among DGI cases throughout the surveillance period. Reviewing the strain’s NCBI Pathogen Detection cluster (PDS000214546.3) at the end of the surveillance period showed that five additional genomes from nationwide genomic surveillance had been added to the cluster, with their specimen collection dates ranging from May 2024 to January 2025.

Guided by findings from genomic analyses, epidemiologists completed investigations of DGI cases per standard protocols including patient interview and medical record review. Of the 13 ST16676-infected cases interviewed, 12 (92.3%) resided within the Minneapolis-St. Paul-Bloomington metropolitan area, 11 (84.6%) were male, and 9 (69.2%) were aged 15 to 44 years. Seven (53.8%) reported anonymous sexual encounters with multiple partners, three (23.1%) of which reported substance use while doing so. Two (15.4%) reported having used doxycycline post-exposure prophylaxis (DoxyPEP). Nine (69.2%) reported histories of prior sexually transmitted infections (STIs), of which 4 (44.4%) reported gonorrhea and 5 (55.5%) reported HIV. Epidemiologic investigation confirmed a direct link between two cases, whose isolates’ genomes were genetically identical at 0 SNPs. The fourteenth case, who refused interviews, had an isolate that was identical at 0 SNPs to the two directly linked isolates, lived in an adjacent state, and received care for the infection in Minnesota.

Phenotypic antimicrobial susceptibility test (AST) results were available in medical records of 10 (71.4%) of the 14 ST16676-infected patients. Consistent with results from NG-STAR and AMRFinderPlus analyses, all 10 isolates showed phenotypic resistance to tetracycline and ciprofloxacin. All 10 isolates exhibited phenotypic susceptibility to ceftriaxone.

## CONCLUSIONS

These findings highlight the importance of DGI surveillance and the utility of genomic surveillance for STIs. Prompt case investigations spurred by genomic analysis allowed epidemiologists to identify a direct link between DGI cases and notify a neighboring state health agency of transmission. Phylodynamic approaches also yielded insights into rates at which DGI-associated strains can emerge, by estimating a timeline of weeks to months between the estimated T_MRCA_ of a strain and the presentation of infected cases at healthcare facilities.

Prospective WGS detected the emergence of a tetracycline-resistant strain of *N. gonorrhoeae* that replaced the predominant DGI-associated strain from the previous year. The sudden emergence of a DGI-associated strain that carries both a *porB1a* allele and a tetracycline resistance gene on a mobilizable plasmid poses significant epidemiological concern, given the use of doxycycline post-exposure prophylaxis (DoxyPEP) to prevent transmission of gonorrhea [10, 15]. Additionally, the presence of two AMR genes on separately mobilizable plasmids highlights the importance of monitoring horizontal gene transfer in genomic surveillance of *N. gonorrhoeae*. Continuing prospective genomic surveillance including performing large-scale studies of the evolution of DGI-causing *N. gonorrhoeae* strains will help the field more thoroughly understand and intervene against this public health threat.

## Data Availability

Sequencing reads and genome assemblies for the N. gonorrhoeae genomes sequenced for this study are publicly available on the National Center for Biotechnology (NCBI) website under BioProject number PRJNA1204341.

https://www.ncbi.nlm.nih.gov/bioproject/1204341

## ACKNOWLEDGEMENTS

Funding for this investigation was provided through the following Centers for Disease Control and Prevention (CDC)-funded grants: Strengthening STD Prevention and Control for Health Departments Award NH25PS005172, Emerging Infections Program NU50CK000648, Epidemiology and Laboratory Capacity NU51CK000361 and NU50CK000508, and Pathogen Genomics Centers of Excellence NU50CK000628. These findings do not necessarily reflect the official opinions of the agencies that funded this work.

We acknowledge Amber Poppe of Red Door Clinic for her support of this epidemiological investigation. We also acknowledge Marcie Babcock, Jeffrey Dennis, Hannah Friedlander, Brian Kendrick, Dakota Schneider, and Jennifer Zipprich of the Minnesota Department of Health for their support of STI surveillance in Minnesota. We acknowledge John C. Cartee and Sandeep J. Joseph for conducting nationwide genomic surveillance that contextualized the results of our local investigations. We also thank the NCBI Pathogen Detection team, as well as all healthcare providers in Minnesota and Wisconsin who provided clinical care to patients with DGI and reported cases to the Minnesota Department of Health.

Sequencing reads and genome assemblies for the *N. gonorrhoeae* genomes sequenced for this study are publicly available on the NCBI website under BioProject number PRJNA1204341.

## APPENDIX

### METHODS

#### Single nucleotide polymorphism analysis of ST16676 genomes

Consistent with the standard Minnesota Department of Health bioinformatics protocol for genomic investigations healthcare-associated bacterial infections, we used the Dryad v3.0.0 pipeline to generate reference-based single nucleotide polymorphism (SNP) matrices of ST16676 genomes [1]. Dryad v3.0.0 uses paired short read files as inputs, performs quality control of sequencing reads using FastQC v0.11.8 [2], matches reads to microbial taxonomy using Kraken2 v2.0.8 [3], assembles genomes using Shovill v1.1.0 [4], assesses assembly quality using QUAST v5.0.2 [5], and calls SNPs among genome assemblies based on a provided reference genome using CFSAN SNP Pipeline v2.0.2 [6].

#### Phylodynamic analysis of ST16676 genomes

We constructed a core genome alignment and phylogenetic tree of all ST16676 genomes using Bakta v1.9.4, Panaroo v1.5.0, and IQTree2 v2.3.6, as described in the main text [7–9]. After rerooting the tree to its midpoint, we performed 32 iterations of the TreeTime v0.11.4 algorithm with the rerooted tree, core genome phylogeny, and documented specimen collection dates [10]. These iterations covered all permutations of five binary parameters: whether (1) a clock rate was calculated by default settings or assigned by a pre-calculation step (“treetime clock” command, “--clock-rate” and “--clock-std-dev” flags), (2) the input tree for the iteration was the one generated by the Dryad pipeline or the version of it optimized when pre-calculating clock rates, (3) TreeTime was permitted to reroot the input tree during the iteration (“--keep-root” flag), (4) divergence times were calculated using jointly or marginally most likely inference settings (“--time-marginal” flag), and (4) polytomies were resolved greedily or stochastically (“--stochastic-resolve” flag). All 32 iterations allowed for phylogenetic covariance when estimating T_MRCA_s (“--covariation” flag), calculated 90% confidence intervals of mugration inferences (“--confidence” flag), and reported correlation coefficients of linear molecular clock models. We assessed the convergence of estimated T_MRCA_s in phylodynamic trees by comparing date ranges of 90% confidence intervals for each TreeTime iteration. Specimen collection dates were input as precisely as available to authors; all Minnesota ST16676 genomes were input to the day, whereas contextual genomes downloaded from NCBI were input to the month.

## FIGURES

**Appendix Figure 1:**
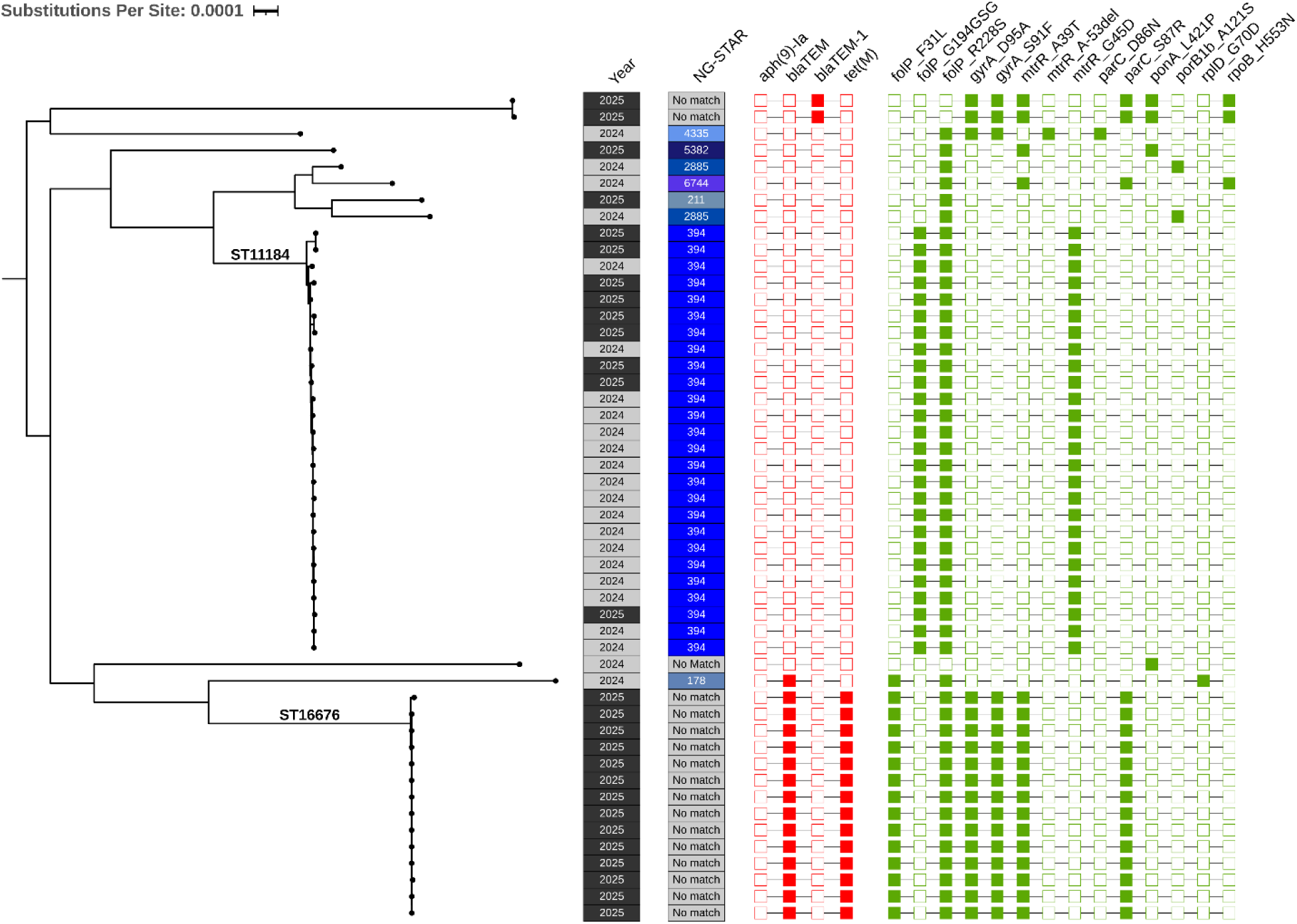
Midpoint-rooted phylogenetic tree constructed from an alignment of 1626 core genes shared by 50 genomes of *N. gonorrhoeae* isolates from disseminated infections in Minnesota in 2024 and 2025. Annotations denote calendar year of specimen collection, NG-STAR, and presence or antimicrobial resistance (AMR) markers detected by AMRFinderPlus. AMR markers shown in red denote acquired genes, and those shown in green denote point mutations. This figure was constructed using ITOL software (https://itol.embl.de/).

**Appendix Figure 2:**
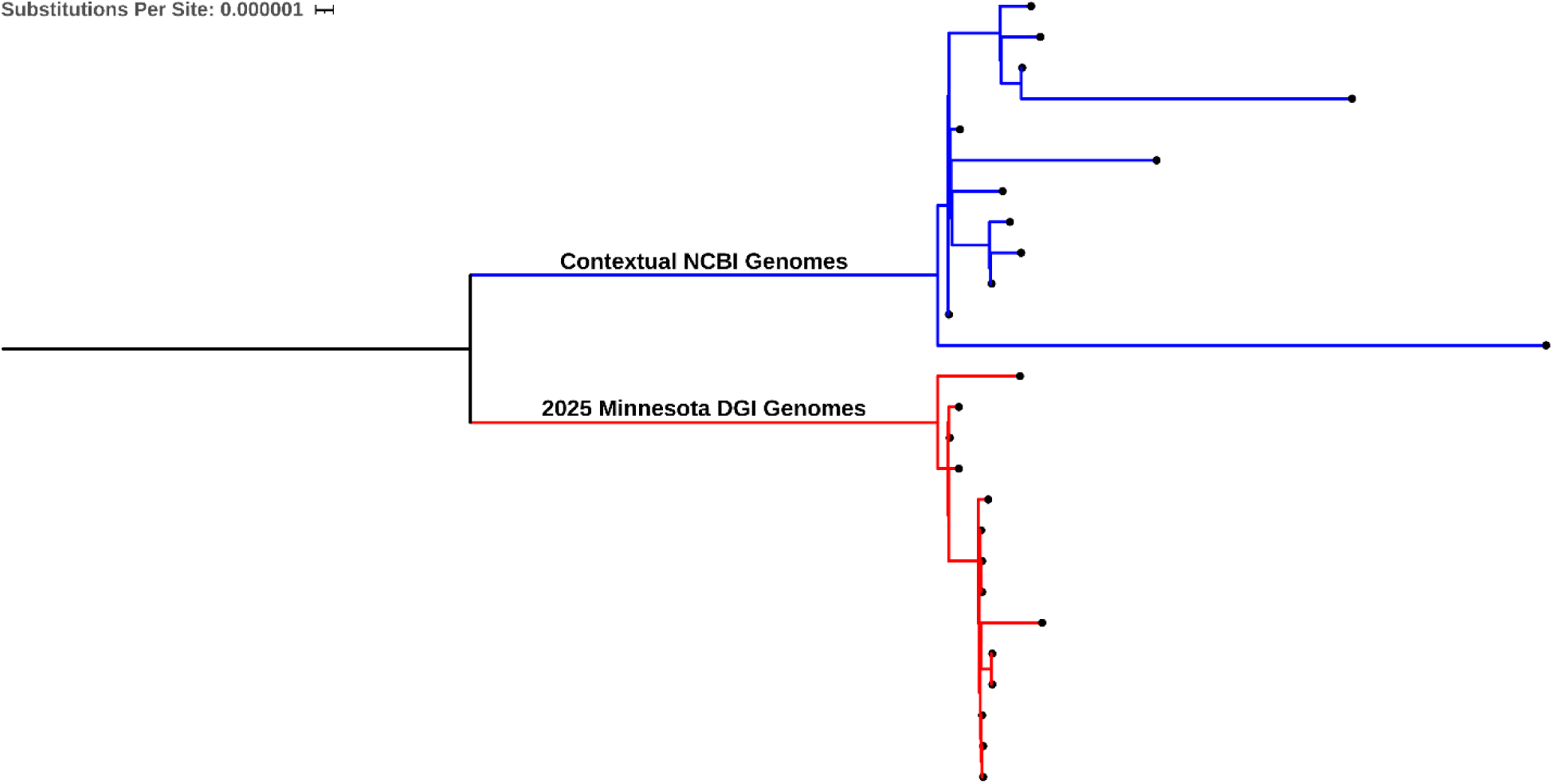
Midpoint-rooted phylogenetic tree constructed from an alignment of 2042 core genes shared among ST16676 genomes. This figure was constructed using ITOL software (https://itol.embl.de/).

